# Affective neural signatures do not distinguish women with emotion dysregulation from healthy controls: A mega-analysis across three task-based fMRI studies

**DOI:** 10.1101/2021.02.03.21251077

**Authors:** M. Sicorello, J. Herzog, T.D. Wager, G. Ende, M. Müller-Engelmann, S.C. Herpertz, M. Bohus, C. Schmahl, C. Paret, I. Niedtfeld

## Abstract

Pathophysiological models are urgently needed for personalized treatments of mental disorders. However, most potential neural markers for psychopathology are limited by low interpretability, prohibiting reverse inference from brain measures to clinical symptoms and traits. Neural signatures—i.e. multivariate brain-patterns trained to be both sensitive *and* specific to a construct of interest—might alleviate this problem, but are rarely applied to mental disorders. We tested whether previously developed neural signatures for negative affect and discrete emotions distinguish between healthy individuals and those with mental disorders characterized by emotion dysregulation, i.e. Borderline Personality Disorder (BPD) and complex Post-traumatic Stress Disorder (cPTSD). In three different fMRI studies, a total sample of 192 women (49 BPD, 62 cPTSD, 81 healthy controls) were shown pictures of scenes with negative or neutral content. Based on pathophysiological models, we hypothesized higher negative and lower positive reactivity of neural emotion signatures in participants with emotion dysregulation. The expression of neural signatures differed strongly between neutral and negative pictures (average Cohen’s *d* = 1.17). Nevertheless, a mega-analysis on individual participant data showed no differences in the reactivity of neural signatures between participants with and without emotion dysregulation. Confidence intervals ruled out even small effect sizes in the hypothesized direction and were further supported by Bayes factors. Overall, these results support the validity of neural signatures for emotional states during fMRI tasks, but raise important questions concerning their link to individual differences in emotion dysregulation.

## 1. Introduction

### 1.1 Background

About 30% of the global population are estimated to suffer from a mental disorder during their lifetime, accompanied by significant human and societal costs (Steel et al., 2014; Whiteford et al., 2013). As for most physical maladies, biological explanations have a long history in this realm (Barondes, 1990). In the last 20 years, functional neuroimaging in particular has become a fundamental research strategy to improve our understanding of mental disorders. Most commonly, clinical researchers, practitioners, and patients are interested in features of the brain to infer clinical traits on a psychological level. For such *reverse inference*, neurobiological features must be both sensitive and specific, i.e. highly predictive of the psychological concept of interest, but not other distinct concepts (Poldrack, 2011). Unfortunately, with few exceptions, classic neural measures like average regional activity are not task-specific (Yarkoni et al., 2011) and have low test-retest reliability (Elliott et al., 2020), precluding reverse inference from brain activity to complex psychological constructs.

*Neural signatures* have been proposed as a solution to this problem (Woo et al., 2017). They can be defined as statistical models, which predict a psychological concept from brain data with great precision, but also distinguish it from similar but meaningfully different concepts (Kragel et al., 2018). For example, a machine learning-based multivariate neural signature of physical pain can be highly predictive of self-reported pain ratings, but distinguishes it from the concept of socio-emotional ‘pain’ following social rejection and vice versa (Woo et al., 2014). Hence, neural signatures ensure interpretability regarding psychological states above other brain-based approaches. Moreover, they might remedy the very low test-retest reliability of non-pattern brain measures (Gianaros et al., 2020; Kragel et al., 2020) as well as increase statistical power by limiting the number of statistical comparisons to a single neural indicator for the process of interest. Despite these advantages, validated neural signatures have rarely been applied to explain individual differences, particularly regarding clinical research questions on mental disorders.

Some mental disorders such as Borderline Personality Disorder (BPD) and complex Post-traumatic Stress Disorder (cPTSD) are characterized by pervasive emotion dysregulation, comprising increased emotional reactivity and deficits in emotion regulation (American Psychiatric Association, 2013; Brewin et al., 2017; Carpenter & Trull, 2013; Linehan, 1993). For the reactivity component, dominant pathophysiological models posit that presumably emotion-generating brain regions are hyperactive in response to negative (or even neutral) stimuli (Brendel et al., 2005; Sicorello & Schmahl, 2020; Swartz et al., 2015). Especially for the amygdala, there is compelling evidence of hyperactivity in these disorders (Bryant et al., 2020; Schulze et al., 2019). Still, amygdala hyperactivity does not warrant reverse inference to heightened emotional reactivity, as it is not specific to negative emotions, but rather involved in a large spectrum of both valence-independent emotional and non-emotional processes (Cunningham & Brosch, 2012; Lindquist et al., 2016; Ousdal et al., 2008; Sander et al., 2003; Todorov, 2012; Wager et al., 2015). Hence, there is still no clear evidence demonstrating emotional hyperreactivity on a brain basis in these disorders.

Several neural signatures of emotions have been developed which are suitable to address this issue, which draw from sparse distributed information across the brain. The picture induced negative emotion signature (PINES; Chang et al., 2015) predicted one-item self-ratings of negative affect following negative pictures with a product-moment correlation above .90, outperforming single resting-state networks and regions, demonstrated dissociability from neural patterns of physical pain, and maintained its cross-validated accuracy in a hold-out sample. Complementary to this pattern for global negative affect, Kragel and LaBar (2015) developed seven patterns which distinguish discrete video-induced emotions from each other at an accuracy close to 40% (chance is ≈14%), including the emotions of fear, anger, sadness, surprise, amusement, contentment and a neutral reference state. Classification accuracy was also above chance when tested on music clips, supporting cross-modal validity. Moreover, in a large resting state fMRI sample of young healthy university students, spontaneous activity of the sadness pattern was associated with an epidemiological depression scale, while the fear pattern was associated with trait anxiety (Kragel et al., 2016). This study provides first evidence that individual differences in the expression of neural emotion networks might map on traits related to the differential experience of emotions on a self-report level.

Expanding this approach to a clinical setting, we tested herein whether the activity of these previously developed neural signatures for general negative affect (i.e. PINES; Chang et al., 2015) and discrete emotions (Kragel & LaBar, 2015) in response to pictures of negative (versus neutral) scenes distinguished women with emotion dysregulation from healthy controls. Negative scenes are among the most common stimuli to study negative emotional reactivity in mental disorders (McDermott et al., 2018). Analyses were conducted across three datasets, each including a clinical group characterized by emotion dysregulation (2 BPD, 1 cPTSD), aggregating results with a mega-analytic approach based on individual participant data.

First, we tested whether neural signatures were differentially expressed in the two experimental conditions. When viewing negative pictures, we expected the pattern expression of negative affect (PINES signature) as well as fear, anger, and sadness (discrete emotion signatures) to be increased (hypothesis 1). Second, for the main research question, common models of the disorders predict heightened reactivity of negative emotions. Here, this translates to increased reactivity of the patterns for negative affect as well as fear, anger, and sadness in participants with emotion dysregulation (hypothesis 2).

Previously, we observed that naturalistic everyday life stressors are associated not only with higher negative affect, but also lower positive affect (Sicorello, Dieckmann, et al., 2020). Therefore, we included additional analyses on neural signatures for positive emotions as well. We predicted the pattern expression of amusement and contentment to be decreased in the negative condition. We predicted stronger deactivation of these patterns in the emotion dysregulation groups. For the surprise pattern, we expected a higher expression in the negative condition, but had no directional between-group hypothesis. Last, the neutral pattern indicates the presence (or absence) of any discrete emotional state. As the paradigm is designed to elicit negative emotions, we expected neutral states to be decreased in the negative condition and more strongly so in the emotion dysregulation group.

## 2. Methods and Materials

### 2.1 Samples and Procedure

Three studies comprising a total of 192 women were included in the analyses of which 111 had a diagnosis of BPD or cPTSD. All participants were presented negative and neutral pictures during fMRI.

Study 1 comprised 57 women (29 with BPD, 28 healthy controls) who participated in a randomized controlled trial on BPD psychotherapy (German Clinical Trials Register: DRKS00000778). Only results from cross-sectional data collected before the intervention are reported here. Participants completed an fMRI experiment with three event-related runs, all with the same structure and number of trials. Each run involved a negative and a neutral condition presented after a “view” instruction. Either negative pictures or pictures of objects where shown, respectively. The experiment also involved regulate-conditions that were not analyzed here, where participants had to regulate their emotional response. Pictures were presented for 6s. Longitudinal results on therapy-effects in this sample have been published previously (Niedtfeld et al., 2017; Schmitt et al., 2016; Winter et al., 2017).

Study 2 comprised 40 women (20 with BPD, 20 healthy controls), who completed three runs of a picture viewing task with different designs: block-design (one picture per block, 18s), mixed-design (three pictures per block, 6s each), and event-related design (6s per picture). Participants viewed negative pictures (negative condition) and scrambled images (neutral condition). Data on the healthy group have been published previously (Paret et al., 2014).

Study 3 comprised 95 women (62 with cPTSD, 33 healthy controls), who were recruited from a larger randomized controlled psychotherapeutic trial (German Clinical Trials Register: DRKS00005578), and therapy-effects were recently published (Bohus et al., 2020). Only results from cross-sectional data collected before the intervention are reported here. Additionally to the DSM-5 criteria for PTSD, participants met at least three out of nine DSM-IV criteria for BPD, including criterion six for emotional instability. Negative pictures and neutral pictures were presented as distractors within a Sternberg working memory task for 1.5s and entered the analysis as negative condition and neutral condition, respectively. Neutral pictures matched with the negative pictures for complexity and content were used in the neutral baseline condition. FMRI data from 34 women of the cPTSD group have been published previously to test a different hypothesis against a trauma-exposed healthy control group (Sicorello, Thome, et al., 2020). The trauma-exposed control group was not included in the analyses here.

Comprehensive descriptions of sample characteristics, designs, procedures, scanning parameters, and preprocessing for all three studies can be found in the supplemental material.

### 2.2 Pattern expression

We downloaded the pattern-masks of each neural signature (PINES and the seven discrete emotion signatures) from the CANlab github repository: https://github.com/canlab. These pattern masks are freely available and consist of a brain image with a regression weight for each brain voxel. Pattern expression was calculated as the dot product between the pattern mask and an image containing beta weights from the first-level analysis for the respective regressor of interest (negative or neutral condition), separately for each picture condition, run, and participant. For the PINES, pattern expression reflects the predicted negative affect rating. For discrete emotions, pattern expression is a continuous indicator to what degree a given emotion category is more likely than the remaining categories. Notably, expression values cannot be directly compared between studies, as their scale depends on scanning parameters, scanner-specific gain and signal characteristics, and analysis choices. Expression values can, however, be compared across task conditions and participants if these values can be assumed to be constant across participants. As an index of reactivity, pattern expression during the neutral condition was subtracted from pattern expression during the negative condition.

As an indicator of internal consistency, we calculated the reliability for the pattern responses as Cronbach’s alpha between experimental runs when more than one run was available (studies 1 and 2). All runs occurred in the same fMRI session. For study 1, pattern responses had a mean reliability of α = .58, ranging from α = .48 for anger to α = .66 for the PINES and fear. As could be expected from previous reports (Gianaros et al., 2020; Kragel et al., 2020), the reliability was higher for pattern expression than for the mean response in an amygdala-hippocampal region-of-interest (ROI; α = .14), which was defined from the thresholded mask of a previous functional meta-analysis on emotion processing in BPD, (Schulze et al., 2019; https://identifiers.org/neurovault.collection:3751). For study 2, pattern responses had a mean reliability of α = .64, ranging from α = .56 for amused to α = .72 for fear. Again, reliability of the amygdala-hippocampal ROI was substantially lower at α = .31. The correlation between pattern expressions in the event-related design and the two block designs was lower than between the two block designs, but not in a range indicating conclusive differences, given the sample size: *r*(event-related, block) = .26, *r*(event-related, mixed-block) = .37, *r*(block, mixed-block) = .57.

### 2.3 Statistical Analyses

#### 2.3.1 Negative versus neutral condition

To test whether the expression of neural signatures differed between the negative and the neutral condition in studies 1-3, reflecting pattern reactivity, one-sample *t*-tests were conducted on the difference scores. Cohen’s *d* was calculated as the mean difference score divided by the standard deviation of difference scores. The three runs of study 1 were averaged for this analysis, as the runs showed good compatibility in terms of sufficient internal consistency and only small differences in mean effects. Runs of study 2 were analyzed separately, to allow the inspection of design-dependent effects and as the three runs had large differences in mean activations, due to the different stimulus presentation parameters.

The corresponding within-person mega-analysis was conducted using a two-level multilevel analysis framework, with difference scores nested within participants (because of the multiple runs in studies 1 and 2). The difference score Δ_ijk_ of run *i* within participant *j* of study *k* was regressed on a fixed intercept γ_000_, including random intercepts for study-participants ζ_0jk_. as well as a residual term ε_ijk_: Δ_ijk_ = γ_000_ + ζ_0jk_ + ε_ijk_. Due to the low number of studies, the study-wise random intercept ζ_00k_ was not included. Moreover, Δ_ijk_ was scaled on the run-specific standard deviation *SD*_i·k_. With this scaling, γ_000_ is in the metric of the Cohen’s *d* used for single study analyses and on a compatible scale between studies and runs, regardless of e.g. design effects. All frequentist multilevel analyses were conducted using the lmer function of the lme4 package in R version 4.0.3 (Bates et al., 2015) and restricted maximum likelihood estimation.

#### 2.3.2 Group effects

For single studies, differences between the clinical and the healthy groups were tested with two-sample *t*-tests for unequal variances and pattern reactivity (Δ) as the dependent variable. Cohen’s *d* was calculated as the difference in group means divided by the pooled standard deviation.

The mega-analysis was specified as Δ_ijk_ = γ_100_(group) + ζ_0jk_ + ε_ijk_, where γ_100_ represents the fixed effect of group. As for within-analysis, the corresponding random effect for group ζ_10k_(group) was not included due to the low number of studies. The group variable was recoded within runs, so that all intercepts (and their variance) are zero. Therefore, the fixed intercept γ_000_ and its variance between studies ζ_00k_ can be omitted from the model. For balanced group sizes (study 2), this can be achieved by coding groups as -0.5 and 0.5, with the regression weight representing the mean difference between groups. For unbalanced group sizes (studies 1 and 3), weighted effect coding was used (Grotenhuis et al., 2017). Moreover, Δ_ijk_ was standardized within runs by subtracting the run-specific mean and dividing by the run-specific pooled standard deviation. With this standardization, γ_100_ is in the metric of Cohen’s *d*, as used for single study analyses.

#### 2.3.3 Bayes factors

Bayes factors were calculated for all models to quantify the relative evidence of the *H*_*0*_ over the *H*_*1*_ (e.g. effect = 0 versus effect ≠ 0), using the low information cauchy prior with a scale factor of 0.707, which is the default of the R package used here and was previously suggested for psychological applications (Wagenmakers et al., 2018). Bayes factors are a ratio between p(Data|*H*_*1*_) and p(Data|*H*_*0*_), with values above 3 (or below 1/3) often used as a minimum cutoff for claims of evidence in favor of one hypothesis over the other, although continuous interpretations are recommended as well (Jarosz & Wiley, 2014). *BF*_*10*_ denotes evidence for the *H*_*1*_, divided by the evidence for *H*_*0*_; *BF*_*01*_ denotes evidence for the *H*_*0*_, divided by the evidence for *H*_*1*_. *BF*_*10*_ equals 1/*BF*_*01*_ and vice versa.

To compute Bayes factor for tests in singles studies, the function ttestBF() of the Bayes factor package was used in R (Morey & Rouder, 2018). For mega-analyses, the multilevel models were refitted using the *brms* package (Bürkner, 2017), comparing models with (*H*_*1*_) and models without (*H*_*0*_) the effect of interest using the function bayes_factor().

In accordance with our hypotheses stated in the introduction, all Bayes factors reflected directional one-sided tests, except for the between-group effect of surprise. This was achieved by modelling a half-cauchy for the *H*_*1*_ in the hypothesized direction. We argue this is appropriate here, as the Bayes factor should reflect evidence for/against the alternative hypothesis of interest, e.g. neural expression of fear is higher when viewing pictures with negative content (and neither zero *nor* lower).

#### 2.3.4 Reproducible Analyses

Data and annotated R scripts to reproduce the main analyses can be found on: https://github.com/MaurizioSicorello/MVPAemoDys_Analyses.git.

Demographic information used for sample description and fMRI images are not openly provided. Requests for primary data should be addressed directly to the corresponding author.

## 3. Results

### 3.1 Comparison between negative and neutral pictures

In line with our hypothesis, both mega-analyses and single-study analyses indicated that neural signatures of negative affect and negative emotions were expressed more strongly while viewing negative pictures, except for the sadness pattern (figure 1, table 1). Likewise, neural signatures of positive emotions were expressed more strongly in the neutral conditions. Effect sizes were overall large, ranging between *d* = 0.81 (anger) and *d* = 2.07 (PINES/negative affect). Only the signature for sadness had a small effect of *d* = -0.23, which went in the opposite direction than expected, i.e. sadness was expressed more strongly in the neutral condition. The null hypothesis that the condition effect for sadness is zero or negative was 60 times more likely than the hypothesized positive effect, i.e. increased neural expression in the negative condition. Study 2 indicated that mixed-block design elicited the largest effects and the event-related design the smallest effects, as has been previously reported for the mass univariate ROI approach (Paret et al., 2014).

**Table 1:**
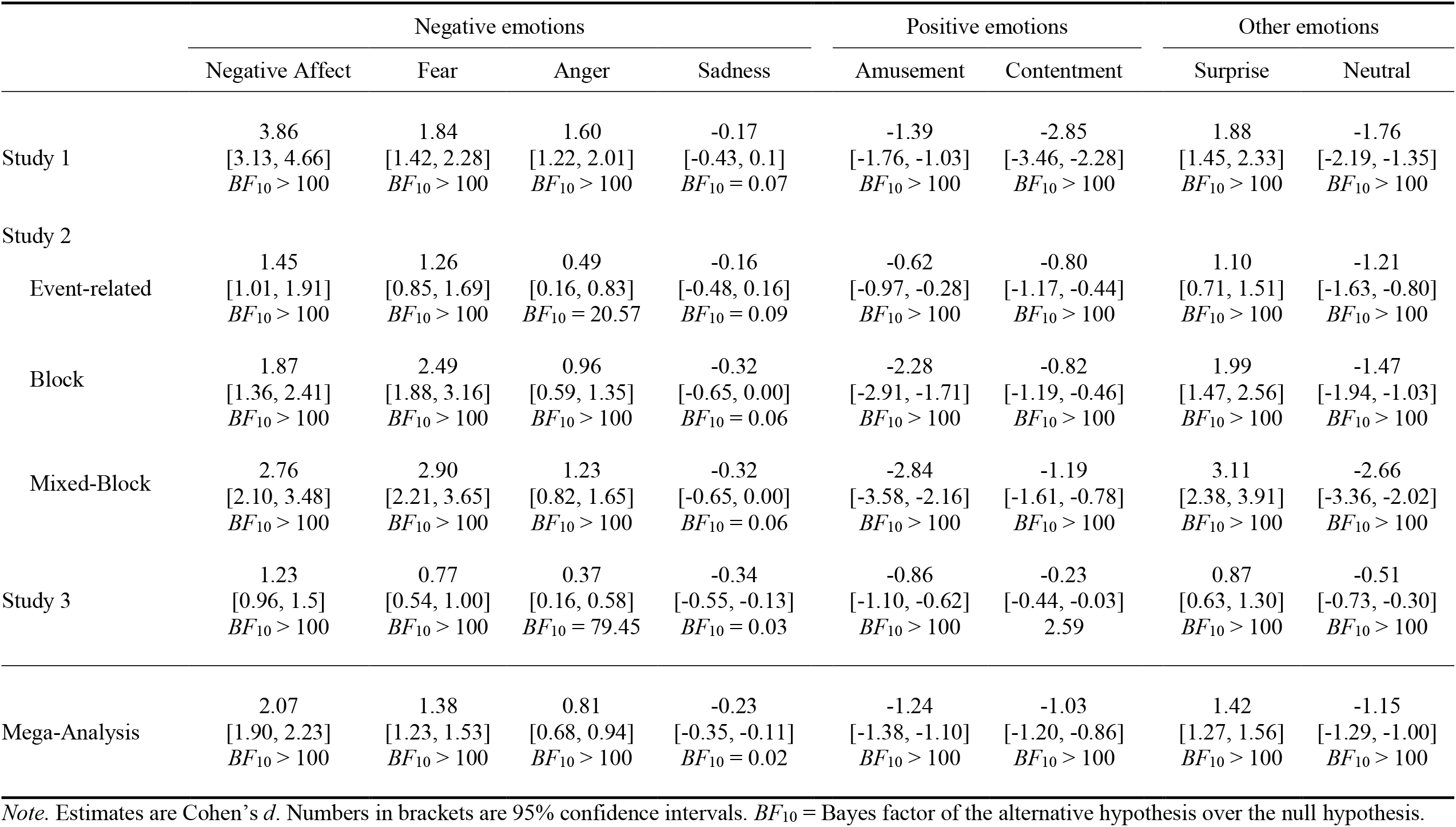
Differences in neural pattern expression between negative and neutral condition

**Figure 1.**
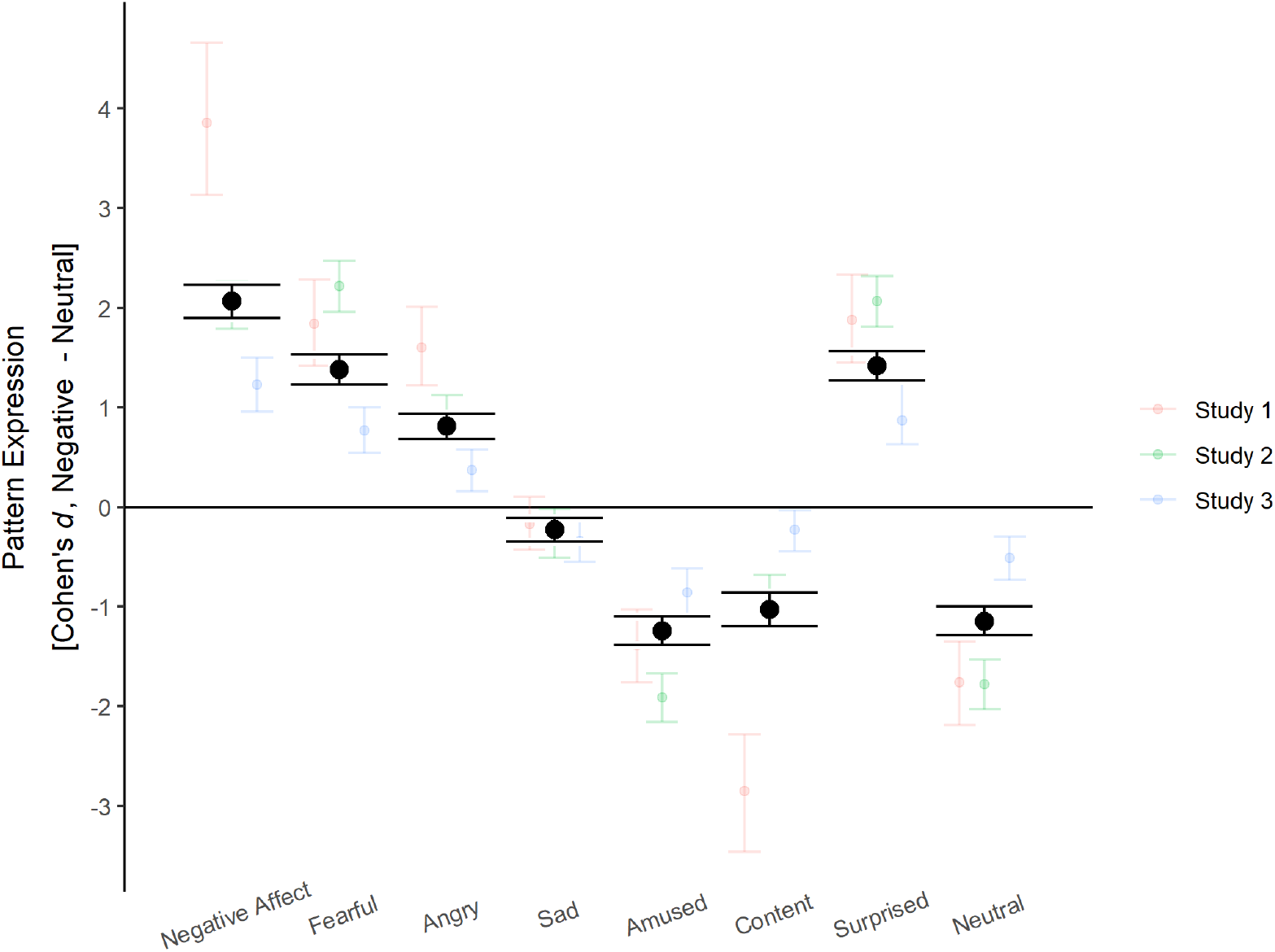
Differences in the expression of neural emotion signatures between the negative and the neutral condition. Error bars show 95% confidence intervals.

In the original validation study, the PINES distinguished the highest and the lowest negative affect ratings at an accuracy of 93.5% (Chang et al., 2015). A logistic regression of picture condition on PINES expression revealed mostly lower but compatible accuracies, with the highest accuracy in the mixed-block design of study 2 and the lowest accuracy in study 3, which had the shortest stimulus presentation duration: Study 1 = 82% [74.21%, 88.94%]; Study 2_Event-Related_ = 71.25% [60.05%, 80.82%]; Study 2_Block_ = 87.50% [78.21%, 93.84%]: Study 2_Mixed_ = 97.50% [91.26%, 99.70%]; Study 3 = 64% [56.41%, 70.52%] (95% confidence intervals in brackets calculated based on Clopper & Pearson (1934).

In sum, these results overall support our first hypothesis that neural signatures of emotions are differentially expressed when viewing negative and neutral pictures in the hypothesized directions, except for the sadness pattern. The estimated effect sizes were very large, but also appeared to depend on design aspects of the studies.

### 3.2 Comparison between clinical groups and healthy controls

Most mega-analytic group effects were very small (all |*d*| ≤ 0.17; figure 2, table 2). Contrary to hypothesis 2—i.e. higher neural pattern reactivity of negative emotions in participants with emotion dysregulation compared to healthy controls—the former actually showed lower reactivity of neural signatures for negative affect, fear, and anger. The upper confidence limit for these three emotions did not include values higher than *d* = 0.12 and Bayes factors favored the null hypothesis of equal or smaller neural signature reactivity in the emotion dysregulation groups. While the emotion dysregulation group did show the expected tendency of higher expression for sadness, the effect was very small (*d* = 0.06), confidence intervals covered zero and had an upper limit at a small effect size of *d* = 0.31, and the Bayes factor favored the null (*BF*_*01*_ = 9.54). Moreover, the condition-wise analyses indicated this emotion signature might not be a valid measure given the stimulus material. Group effects for neutral states, amusement, contentment, and surprise did not differ considerably from zero. These results were supported by Bayes factors, except for surprise, whose Bayes factor was relatively inconclusive (*BF*_*01*_ = 2.15). On a single study-basis, this pattern was overall present in studies 1 and 2. The descriptive effect directions in study 3 were more compatible with the theoretical predictions, albeit with miniscule effect sizes and inconclusive Bayes factors. These results were stable when a binary indicator for psychotropic medication was included as a covariate (figure S1).

**Table 2:** Differences in neural pattern reactivity (negative – neutral condition) between emotion dysregulation and healthy control group

|  | Negative emotions |  |  |  | Positive emotions |  | Other emotions |  |
| --- | --- | --- | --- | --- | --- | --- | --- | --- |
|  | Negative Affect | Fear | Anger | Sadness | Amusement | Contentment | Surprise | Neutral |
| Study 1 | -0.09<br>[-0.61, 0.43]<br>$BF_{01} = 4.76$ | 0.01<br>[-0.51, 0.53]<br>$BF_{01} = 3.57$ | -0.45<br>[-0.97, 0.08]<br>$BF_{01} = 9.09$ | 0.13<br>[-0.39, 0.65]<br>$BF_{01} = 2.50$ | -0.45<br>[-0.97, 0.08]<br>$BF_{01} = 0.06$ | 0.49<br>[-0.04, 1.02]<br>$BF_{01} = 10.0$ | 0.16<br>[-0.37, 0.68]<br>$BF_{01} = 3.23$ | 0.29<br>[-0.24, 0.81]<br>$BF_{01} = 7.14$ |
| Study 2 |  |  |  |  |  |  |  |  |
| Event-related | -0.37<br>[-0.99, 0.26]<br>$BF_{01} = 6.25$ | -0.36<br>[-0.98, 0.27]<br>$BF_{01} = 6.25$ | -0.20<br>[-0.82, 0.42]<br>$BF_{01} = 4.76$ | -0.39<br>[-1.01, 0.24]<br>$BF_{01} = 6.25$ | -0.08<br>[-0.70, 0.54]<br>$BF_{01} = 2.70$ | 0.32<br>[-0.31, 0.94]<br>$BF_{01} = 5.88$ | 0.15<br>[-0.47, 0.77]<br>$BF_{01} = 2.94$ | 0.36<br>[-0.27, 0.98]<br>$BF_{01} = 6.25$ |
| Block | -0.99<br>[-1.64, -0.33]<br>$BF_{01} = 11.11$ | -0.95<br>[-1.60, -0.29]<br>$BF_{01} = 11.11$ | -0.34<br>[-0.96, 0.28]<br>$BF_{01} = 5.88$ | 0.30<br>[-0.32, 0.93]<br>$BF_{01} = 1.41$ | 0.68<br>[0.04, 1.31]<br>$BF_{01} = 9.09$ | 0.20<br>[-0.43, 0.82]<br>$BF_{01} = 4.76$ | -0.23<br>[-0.85, 0.39]<br>$BF_{01} = 2.63$ | 0.05<br>[-0.57, 0.67]<br>$BF_{01} = 3.57$ |
| Mixed-Block | -0.37<br>[-0.99, 0.26]<br>$BF_{01} = 6.25$ | -1.11<br>[-1.77, -0.44]<br>$BF_{01} = 12.5$ | -0.21<br>[-0.83, 0.41]<br>$BF_{01} = 5.00$ | 0.18<br>[-0.44, 0.80]<br>$BF_{01} = 2.04$ | 0.51<br>[-0.12, 1.14]<br>$BF_{01} = 7.14$ | 0.17<br>[-0.45, 0.79]<br>$BF_{01} = 4.55$ | -0.01<br>[-0.63, 0.61]<br>$BF_{01} = 3.23$ | 0.12<br>[-0.50, 0.74]<br>$BF_{01} = 4.17$ |
| Study 3 | 0.16<br>[-0.26, 0.58]<br>$BF_{01} = 2.27$ | 0.21<br>[-0.21, 0.63]<br>$BF_{01} = 1.79$ | 0.13<br>[-0.3, 0.55]<br>$BF_{01} = 2.70$ | 0.06<br>[-0.36, 0.48]<br>$BF_{01} = 3.57$ | -0.11<br>[-0.53, 0.32]<br>$BF_{01} = 2.94$ | -0.10<br>[-0.52, 0.33]<br>$BF_{01} = 3.12$ | 0.23<br>[-0.19, 0.65]<br>$BF_{01} = 2.70$ | -0.36<br>[-0.79, 0.06]<br>$BF_{01} = 0.69$ |
| Mega-Analysis | -0.13<br>[-0.38, 0.11]<br>$BF_{01} = 11.56$ | -0.13<br>[-0.39, 0.12]<br>$BF_{01} = 7.28$ | -0.16<br>[-0.39, 0.08]<br>$BF_{01} = 53.01$ | 0.06<br>[-0.18, 0.31]<br>$BF_{01} = 9.54$ | -0.07<br>[-0.32, 0.17]<br>$BF_{01} = 4.79$ | 0.17<br>[-0.08, 0.41]<br>$BF_{01} = 9.68$ | 0.12<br>[-0.13, 0.37]<br>$BF_{01} = 2.15$ | 0.01<br>[-0.24, 0.25]<br>$BF_{01} = 21.70$ |

*Note.* Estimates are Cohen's $d$ . Numbers in brackets are 95% confidence intervals. $BF_{01}$ = Bayes factor of the null hypothesis over the alternative hypothesis.

**Figure 2.**
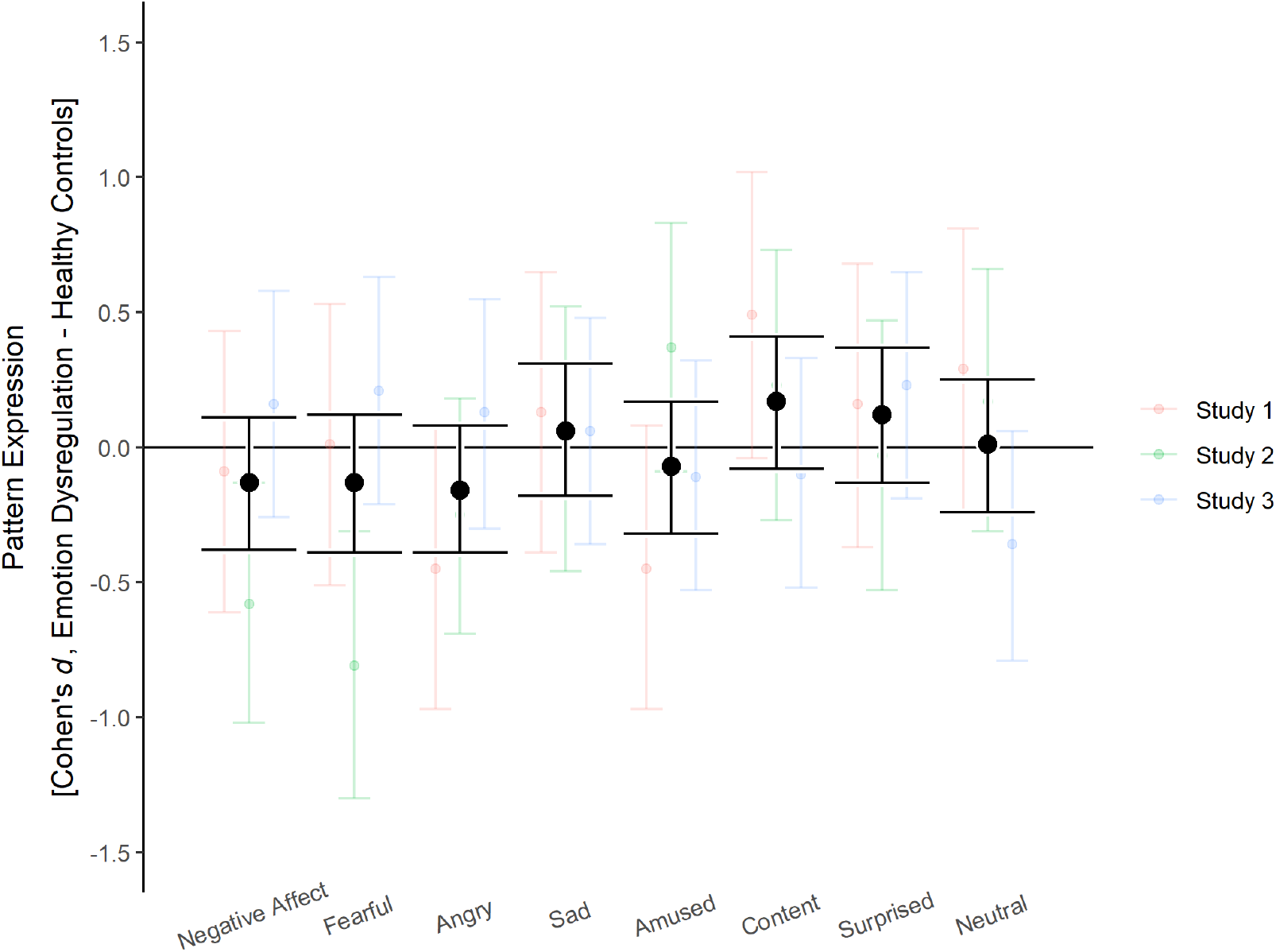
Group differences in the reactivity of neural emotion signatures between participants with emotion dysregulation and healthy controls. Error bars show 95% confidence intervals.

### 3.2 Exploratory Analyses: Group effects on the neutral baseline

There is some evidence that people with emotion dysregulation have a higher propensity to interpret neutral stimuli as negative (Daros et al., 2013; Mitchell et al., 2014), accompanied by heightened amygdala responses (Donegan et al., 2003; Lischke et al., 2017; Niedtfeld et al., 2010). As this might diminish group differences in the negative-neutral contrast, we repeated the between-group analyses of section 3.2 with activation in the neutral condition as the dependent variable, instead of the difference between negative and neutral conditions.

In these analyses, all confidence intervals contained zero by a considerable margin (figure S2). Still, even statistically non-significant group effects on the neutral baseline might diminish group effects on the difference scores used to indicate neural reactivity. In the neutral condition, participants with emotion dysregulation had slightly increased responses for the fear pattern (*d* = 0.17, 95% *CI* = [-0.08, 0.43]) and decreased responses for the contentment pattern (*d* = -0.10, 95% *CI* = [-0.35, 0.15]). Hence, the hypothesized effects for these two patterns might be diminished by group differences in response to the neutral condition. All other effects were in the opposite direction of what would be expected if an increased responsiveness to neutral stimuli accounts for the null effects reported in section 3.2 (e.g. participants with emotion dysregulation had a lower expression of the PINES signature and a higher expression of the neutral signature).

To follow up on the potential attenuation effect for fear and contentment, we repeated the mega-analytic procedure on pattern expression in the negative condition against the implicit baseline (figure S3). The estimates for the fear and contentment patterns were almost perfectly zero, although confidence intervals of the fear pattern still included small to moderate effect sizes (fear: *d* = 0.00, 95% *CI* = [-0.26, 0.26]; contentment: *d* = 0.01, 95% *CI* = [-0.24, 0.26]).

Coincidentally, we observed that the confidence interval of the effect of lower negative affect in the emotion dysregulation group vs. the healthy control group no longer contained zero (*d* = -0.32, 95% *CI* = [-0.57, -0.07]), which differs from the results for the negative-neutral contrast.

## 4. Discussion

To translate neurobiological models of mental disorders into the clinical language of traits and symptoms, neural markers have to be both sensitive *and* specific to the psychological concept of interest. This is rarely the case for properties of discrete anatomical brain regions like the amygdala, which nonetheless has been frequently used as an indicator of negative emotional processes in affect-related disorders, while it is also involved in a broad set of psychological phenomena other than emotions. Here, we used machine learning-based multivariate neural signatures for emotional states to test whether people with emotion dysregulation show signs of hyperreactive neuro-emotional systems. This assumption of leading psychopathological models was assessed in three independent studies from our lab, investigating participants diagnosed with either BPD or cPTSD and healthy controls.

Neural signatures of negative affect (Chang et al., 2015) and discrete emotions (Kragel & LaBar, 2015) showed strong differential expression between the negative and the neutral condition in the expected directions (hypothesis 1), supporting their validity and accuracy, even when transferred to a different lab, experimental design, and population than the initial validation studies. Effect sizes were very large and supported by very large Bayes factors in each of the three studies. Moreover, study 2 indicates that effect sizes might be partly related to stimulus presentation parameters such as exposure time. Notably, the effect observed with the sadness signature was in the opposite direction than expected (neutral > negative condition). As the stimuli were chosen based on valence and arousal ratings, it is possible that sadness- inducing pictures were underrepresented or that sadness is harder to induce with briefly presented pictures.

Most importantly, the neural signatures did not differentiate between participants with and without emotion dysregulation, speaking against the main hypothesis of the present study (hypothesis 2). Except for the sadness and amusement signatures, all effects went in the opposite direction from the theoretical predictions, i.e. smaller negative emotional reactivity and positive emotional reactivity in the emotion dysregulation group vs. the healthy group. The corresponding confidence intervals ruled out even small effect sizes in the expected direction, below |*d*| = 0.20 and Bayes factors favored the null hypothesis for all signatures, except for surprise, which was inconclusive. Similar patterns emerged for separate analyses on studies 1 and 2, while the results in study 3 were less conclusive in terms of Bayes factors. These results could not be explained by a heightened response to the neutral condition in those with BPD and cPTSD, which has been observed previously for amygdala reactivity.

These findings are incompatible with the dominant pathological model of BPD and provide evidence against either the theoretical, experimental, or neurobiological assumptions of the present study, which we discuss below. Either way, important implications arise for future research. To discuss these potential explanations of the reported results, we mainly draw from the BPD literature, as the cPTSD literature is still relatively limited and the BPD criterion for emotional instability was the cardinal criterion for inclusion in study 3.

Showing participants pictures of scenes with negative content is among the most common tasks to experimentally investigate heightened emotional reactivity in mental disorders and affect-related traits. This approach rests on the implicit assumptions that (1) the clinical phenomenon of heightened emotional reactivity is not fully accounted for by more negative environments, a lower threshold for emotional responses, or difficulties in emotion regulation, (2) emotional reactivity can be observed outside of its naturalistic daily life context, and (3) the emotion-inducing effect of experimental stimuli is not limited to stimuli personalized according to thematic relevance. If correct, these assumptions naturally lead to the conclusion that people with emotion dysregulation must have generally hyperresponsive emotion generating biological systems, whose exploration could aid the understanding and treatment of such disorders. Further, our aim to investigate these biological systems with neural signatures was based on the assumption that (4) neural signatures represent the best available neural markers for such systems, due to their high accuracy for emotional states.

Apart from qualitative clinical impression of therapeutic practitioners, there is empirical evidence for increased reactivity to discrete naturalistic everyday life stressors in BPD (Glaser et al., 2008; Hepp et al., 2018). Notably, such studies cannot easily distinguish precisely which aspects of emotion processing are aberrant, due to their relatively low temporal resolution (assumption 1). Experimental settings offer higher control and better temporal resolution, but suffer from limited ecological validity, as stressors are presented outside of their natural context (assumption 2). A recent meta-analytic review found that the literature is surprisingly inconclusive concerning experimentally induced emotional reactions in BPD (Bortolla et al., 2020). While they did find moderate experimental group effects on affective self-ratings in their meta-analysis, many studies did not include a pre-measurement, potentially confounding tonic negative emotions and emotional reactivity, or only had pre- and post-task ratings, which might capture other processes than stimulus-contingent real-time responses. Moreover, peripheral-physiological effects were negligibly small and/or statistically not significant. Interestingly, there was no statistically significant difference in effect sizes dependent on whether stimuli were thematically related to BPD (assumption 3).

Taken together, it is possible that typical laboratory designs, as used in our studies, are not well-suited to probe individual differences in emotional reactivity which generalize to everyday life or that clinical subgroups with opposing phenomenology cancel each other’s effects. Alternatively, it is possible that the neural signatures do not capture the psychological concept of interest well (assumption 4). If the concepts of interest are emotions as they are measured by self-reports, this seems unlikely for the PINES, as it correlated with self-reports above *r* = .90 in both the training and the hold-out sample, which employed a design similar to ours. Still, it is possible that when asked for their mood directly after seeing a negative picture, participants partly rate the picture content, rather than exclusively their emotions, which could have impeded the construct validity of the PINES. Nevertheless, this argument does not hold for the discrete emotion signatures, which distinguish emotion categories and were associated with trait depressiveness and anxiety in a well-powered resting-state study.

Another neurobiological explanation of the null results might be the presence of stable physiological between-person noise (e.g. cerebrovasculature or hematocrit levels; D’Esposito et al., 2003; Yang et al., 2014). A recent meta-analysis demonstrated that test-retest reliability of resting state fMRI diminishes considerably after artefact correction, indicating the presence of such stable between-person noise (Noble et al., 2019). The neural signatures used here have been developed to explain variance without explicit differentiation of the within- or between-person level and their high accuracy might be preferentially due to variance within individuals. Notably, while machine learning-based approaches have been increasingly used to differentiate between clinical groups based on fMRI data (Gao et al., 2018; Woo et al., 2017), these approaches do not necessarily lead to interpretable neural markers, as groups might differ on many confounded dimensions.

### 4.1 Limitations and future directions

The mega-analyses did not include random slopes for studies, as the low number of studies does not allow a sensible estimate of between-study variance. Hence, the generalizability to other experimental investigations is limited and a wider range of effect sizes should be expected (Yarkoni, 2020). This limitation on generalizability is especially important, as studies included only female participants, due to potential gender-differences in symptom presentation (Sansone & Sansone, 2011). Study 2 indicated that stimulus presentation parameters might be one important influence on effect size differences, at least for within-person effects.

Another limitation to consider is the reliability of fMRI-based neural markers (Elliott et al., 2020). Testing the internal consistency for multi-run studies 1 and 2 indicated that reliability was considerably higher for neural signatures than for an amygdala-hippocampal cluster from a BPD meta-analysis, but still lower than desirable, ranging from α = .48 to α = .72. These estimates could be used in future studies to correct expected effect sizes for unreliability in power analyses.

As in most BPD studies which used fMRI designs with negative scenes, there were no affective self-ratings directly following pictures. Such ratings would be necessary to closely replicate the core assumption of the neural emotion signatures, that is, they predict momentary subjective affect ratings by means of BOLD responses to affective stimuli across different populations. More research is urgently needed to confirm the strict validity of neural signatures in clinical populations. Post-session valence ratings of negative pictures did not differ considerably between participants, as has been previously reported (Koenigsberg et al., 2009; Schulze et al., 2011), but are not necessarily a valid surrogate of *momentary* affect, immediately following negative trials. While these tasks have been frequently used, there has been to our knowledge no thorough psychometric validation to ensure their usefulness for research on individual differences on the psychological end. Therefore, we suggest a systematic assessment of their test-retest reliability and validity in terms of associations with clinically relevant traits, independent of neuroimaging techniques. As stated above, it is unclear whether valence ratings following the session should continue to replace self-ratings of affect immediately following image-exposure.

## 4.2 Conclusion

Neural signatures of emotions appear to be valid and transferable tools to investigate within-person relationships, but their utility to understand individual differences remains unclear. Contrary to theoretical expectations, we did not find differences between people with and without emotion dysregulation. We offer to share our analysis pipelines with other research groups to reanalyze existing datasets. This could be done efficiently and lead to a more comprehensive picture of the relationship between neural signatures and emotion-related traits. Apart from neurobiological approaches, more research is needed concerning the psychometric properties and ecological validity of typical experimental tasks used to probe affective traits.

## Data Availability

Data and annotated R scripts to reproduce the main analyses can be found on github.
Demographic information used for sample description and fMRI images are not openly provided. Requests for primary data should be addressed directly to the corresponding author.

https://github.com/MaurizioSicorello/MVPAemoDys_Analyses.git

## Acknowledgements

Thanks are due to Rosemarie Kluetsch, Steffen Hoesterey, Michael Rieß and Claudia Stief for support in data collection, Dorina Winter and Ruth Schmitt for study setup and data processing, to Madita Stirner, Katharina Brunner, and Elena Buck for compiling questionnaire data and to the KFO256-Central Project team for providing excellent organizational support and clinical diagnostics. Study 1 was funded by the German Research Foundation (grant no. SCHM 1526/8- 2; HE2660/7-2). Study 2 was supported by the German Research Foundation (grant no. KFO 256, EN 361/13-2). Study 3 was financed by the German Ministry of Education and Research (BMBF) RELEASE 01KR1303A.

